# Immune escape and attenuated severity associated with the SARS-CoV-2 BA.2.86/JN.1 lineage

**DOI:** 10.1101/2024.04.17.24305964

**Authors:** Joseph A. Lewnard, Parag Mahale, Debbie Malden, Vennis Hong, Bradley K. Ackerson, Bruno J. Lewin, Ruth Link-Gelles, Leora R. Feldstein, Marc Lipsitch, Sara Y. Tartof

**Author notes:** Address for correspondence: Joseph A. Lewnard, Room 5410, 2121 Berkeley Way, Berkeley, California 94720.

## Abstract

The SARS-CoV-2 BA.2.86 lineage, and its sublineage JN.1 in particular, achieved widespread transmission in the US during winter 2023-24. However, the increase in infections was not accompanied by increases in COVID-19 hospitalizations and mortality commensurate with prior waves. To understand shifts in COVID-19 epidemiology associated with JN.1 emergence, we compared characteristics and clinical outcomes of time-matched cases infected with BA.2.86- derived lineages (predominantly representing JN.1) versus co-circulating XBB-derived lineages in December, 2023 and January, 2024. Cases infected with BA.2.86-derived lineages received greater numbers of COVID-19 vaccine doses, including XBB.1.5-targeted and BA.4/BA.5-targeted boosters, in comparison to cases infected with XBB-derived lineages. Additionally, cases infected with BA.2.86-derived lineages experienced greater numbers of documented prior SARS-CoV-2 infections. These associations of BA.2.86-derived lineages with immune escape were confirmed when comparing cases diagnosed during periods when JN.1 was the predominant circulating lineage to cases diagnosed during November, 2023. Cases infected with BA.2.86-derived lineages, or during periods when JN.1 was the predominant circulating lineage, also experienced lower risk of progression to severe clinical outcomes requiring emergency department consultations or hospital admission. Sensitivity analyses suggested under-ascertainment of prior infections, even if differential between cases infected with BA.2.86-derived lineages and non-BA.2.86 lineages, could not explain this apparent attenuation of severity. Our findings implicate escape from immunity acquired from prior vaccination or infection in the emergence of the JN.1 lineage and suggest infections with this lineage are less likely to experience clinically-severe disease. Monitoring of immune escape and clinical severity in emerging SARS-CoV-2 variants remains a priority to inform responses.

## INTRODUCTION

The BA.2.86 SARS-CoV-2 lineage, which is distinguished from the parent BA.2 lineage by over 30 mutations in the spike protein, was detected simultaneously in multiple European countries in July, 2023 (1). A sub lineage (BA.2.86.1.1; “JN.1”) harboring one additional Spike (S) protein mutation (L455S) emerged shortly thereafter and became the dominant circulating lineage in the US by late December, 2023 (2). Similar to other BA.2.86 lineages, JN.1 has been reported to evade neutralizing antibody responses associated with prior SARS-CoV-2 infection and COVID-19 vaccination in comparison to co-circulating lineages derived from XBB.1.5 (3,4). Modification of angiotensin-converting enzyme 2 (ACE2) binding affinity may have further contributed to the establishment of JN.1 and other BA.2.86 lineages (5–7).

While declining rates of clinical SARS-CoV-2 testing prevent comparison of case-based surveillance of JN.1 with earlier phases of the pandemic, detection of SARS-CoV-2 genetic material in wastewater during the JN.1 wave reached levels not seen since the peak of the Omicron BA.1 wave in January, 2021 (8). However, this expansive transmission of JN.1 has not been associated with increases in COVID-19 related hospital admissions or deaths commensurate with the earlier BA.1, BA.4/BA.5, and XBB/XBB.1.5 epidemic waves (9). Assessments of characteristics and clinical outcomes of cases infected with emerging SARS-CoV-2 lineages are needed to interpret whether such epidemiologic observations reflect changes in clinical severity and immune protection (10–15). We therefore compared prior vaccination, documented SARS-CoV-2 infection history, and post-diagnosis healthcare utilization among cases infected with differing lineages within the Kaiser Permanente Southern California (KPSC) healthcare system who were tested in outpatient settings during December, 2023 and January, 2024.

## RESULTS

### Study setting, enrollment and case definitions

The KPSC healthcare system provides managed, integrated care spanning virtual, outpatient, emergency department and inpatient settings to roughly 4.7 million adults residing in southern California, representing roughly 20% of the region’s population. Individuals are enrolled in KPSC plans through employer-sponsored, pre-paid or government-subsidized coverage schemes. Enrolled members closely resemble the general insured population within Southern California (16,17). Electronic health care records capture all in-network care delivery, comprising diagnoses, prescription fills, procedures, laboratory testing, vaccinations, and clinical notes. Records of COVID-19 vaccinations received outside KPSC are imported from the California Immunization Registry (18). Other care delivered out-of-network is ascertained through insurance claim reimbursements, enabling near-complete ascertainment of members’ medical histories.

We conducted a retrospective cohort study leveraging the opportunity for longitudinal follow-up among cases initially diagnosed with SARS-CoV-2 infection in outpatient settings to monitor progression to severe disease outcomes. Our analyses followed cases who had been members of KPSC health plans for ≥1 year from the point of their first documented positive outpatient test between 1 December, 2023 and 30 January, 2024. Over this period, 46,067 eligible individuals tested positive for SARS-CoV-2 in KPSC outpatient settings. Of this population, 7,694 (17%) had tests processed by regional testing laboratories using the TaqPath COVID-19 Combo Kit assay (ThermoFisher Scientific, Waltham, Massachusetts), which provides readout on probes for the S, N, and orf1a/b genes (**Figure 1**; **Table S1**). Dropout of the S gene probe in samples that tested positive for both N and orf1a/b (defined as cycle threshold [c*_T_*] values of ≥37 for S and <37 for N and orf1a/b) provided 98-100% sensitivity and 96% specificity for distinguishing BA.2.86-derived lineages within a sample of 1,078 sequenced specimens from KPSC testing laboratories during the study period (**Table S2**), consistent with observations in other settings (19).

**Figure 1:**
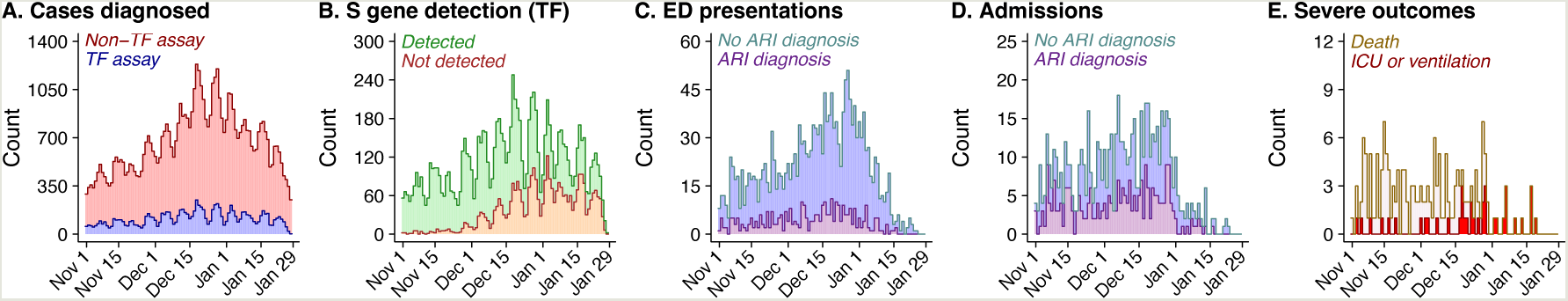
Testing, S-gene targeted taction, and clinical outcomes during the study period. Panels illustrate (**a**) the number of outpatient cases diagnosed daily from tests processed on ThermoFisher TaqPath COVID-19 Combo Kit (TF) assays or non-TF assays; (**b**) the daily frequency of TF-tested specimens yielding positive results with S gene detected (non-BA.2.86 lineages) or S-gene target failure (BA.2.86-derived lineages); (**c**) the daily frequency of outpatient cases with positive SARS-CoV-2 testing results (organized by date of test) who experienced emergency department (ED) presentations within 14 days of testing, stratified according to presence or absence of acute respiratory infection (ARI) diagnoses associated with their ED presentation; (**d**) the daily frequency of outpatient cases with positive SARS-CoV-2 testing results (organized by date of test) who experienced hospital admission within 28 days of testing, stratified according to presence or absence of ARI diagnoses associated with their hospital admission; and (**e**) the daily frequency of outpatient cases with positive SARS-CoV-2 testing results (organized by date of test) who experienced intensive care unit (ICU) admission, initiation of mechanical ventilation, or death within 60 days of testing.

We therefore used S-gene target failure (SGTF) as a proxy for infection with JN.1 or other BA.2.86-derived lineages and defined the primary analytic cohort as the subset of cases whose specimens were processed using TaqPath COVID-19 Combo Kit assays (*N*=7,694). This population closely resembled other outpatient-diagnosed cases at KSPC over the same period in terms of sex, health status, prior-year healthcare utilization, and community socioeconomic characteristics. However, cases tested via TaqPath COVID-19 Combo Kit assays were modestly younger in comparison to other outpatient-diagnosed cases (median age 46 vs. 50 years, respectively) and more racially and ethnically diverse (22% vs 34% identifying as non-Hispanic White, respectively; **Table S1**). Within this primary analytic cohort, cases infected with JN.1 or other BA.2.86-derived lineages (*N*=3,080) did not differ appreciably from those infected with other lineages (*N*=4,614) in terms of age, sex, or racial/ethnic distribution, comorbidity burden, prior-year patterns of healthcare utilization, community socioeconomic characteristics, or receipt of nirmatrelvir-ritonavir (**Table 1**).

**Table 1:**
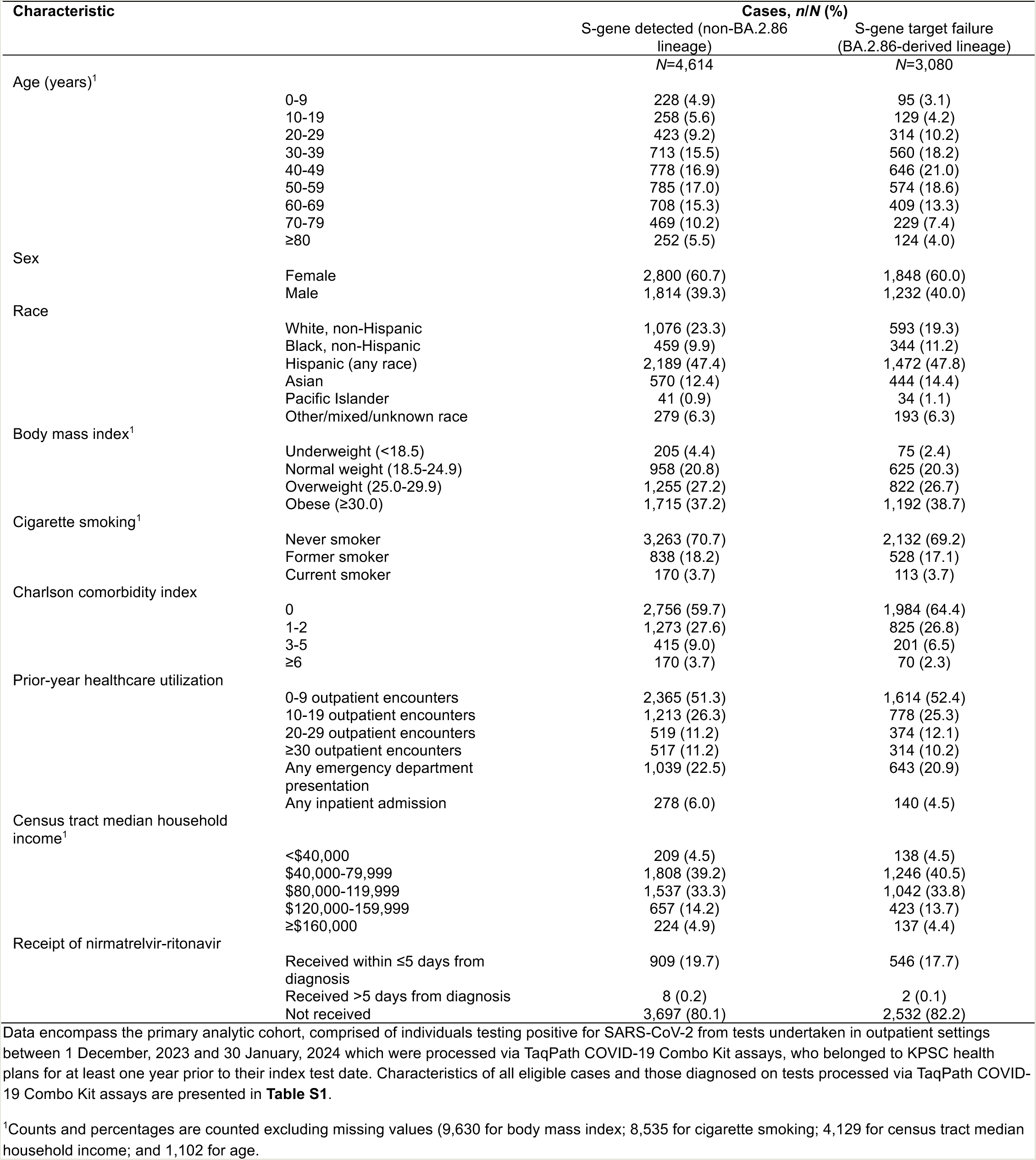
Characteristics of individuals infected with SARS-CoV-2 according to lineage.

### Comparison of immune history by infecting lineage

We first compared vaccination history among cases infected with BA.2.86-derived lineages or non-BA.2.86 lineages, hypothesizing that immune escape by BA.2.86 would lead to detection of related lineages among individuals with a history of COVID-19 vaccination. Consistent with this hypothesis, 272 (9% of 3,080) cases infected with BA.2.86-derived lineages and 569 (12% of 4,614) cases infected with non-BA.2.86 lineages had received no COVID-19 vaccine doses prior to diagnosis (**Table 2**). In conditional logistic regression analyses matched on testing week and controlling for measured characteristics of cases (see **Methods**), adjusted odds of receipt of 5, 6, and ≥7 COVID-19 vaccine doses were 38% (95% confidence interval: 9-74%), 51% (17-95%), and 60% (7-138%) higher among cases infected with BA.2.86-derived lineages in comparison to cases infected with non-BA.2.86 lineages. Cases infected with BA.2.86-derived lineages also had 4-20% higher adjusted odds of having received 1-4 COVID-19 vaccine doses, although the possibility of no difference could not be excluded within some smaller case strata for these lower-dose exposures. Similar patterns persisted in subgroup analyses restricted to cases documented to have experienced ≥1 or ≥2 prior SARS-CoV-2 infections, suggesting relationships between prior vaccination and infecting lineage were not exclusively mediated by antecedent effects of vaccination on cases’ risk of prior infection (**Table S3**).

**Table 2:** Prior vaccination and documented SARS-CoV-2 infection among individuals infected with SARS-CoV-2 according to infecting lineage.

| Exposure |  | Cases, *n/N* (%) |  | Odds ratio (95% CI), JN.1 vs. non-JN.1 infection |  |
| --- | --- | --- | --- | --- | --- |
|  |  | S-gene detected (non-BA.2.86 lineage) | S-gene target failure (BA.2.86-derived lineage) | Unadjusted<sup>1</sup> | Adjusted<sup>2</sup> |
|  |  | *N*=4,614 | *N*=3,080 |  |  |
| Prior vaccination | 0 vaccine doses | 569 (12.3) | 272 (8.8) | ref. | ref. |
|  | 1 vaccine dose | 122 (2.6) | 66 (2.1) | 1.08 (0.83, 1.41) | 1.04 (0.79, 1.36) |
|  | 2 vaccine doses | 901 (19.5) | 564 (18.3) | 1.16 (1.01, 1.34) | 1.10 (0.94, 1.28) |
|  | 3 vaccine doses | 1,453 (31.5) | 1,011 (32.8) | 1.27 (1.11, 1.45) | 1.20 (1.04, 1.39) |
|  | 4 vaccine doses | 824 (17.9) | 570 (18.5) | 1.25 (1.08, 1.44) | 1.20 (0.98, 1.47) |
|  | 5 vaccine doses | 415 (9.0) | 331 (10.7) | 1.34 (1.14, 1.58) | 1.38 (1.09, 1.74) |
|  | 6 vaccine doses | 285 (6.2) | 231 (7.5) | 1.32 (1.11, 1.58) | 1.51 (1.17, 1.95) |
|  | ≥7 vaccine doses | 45 (1.0) | 35 (1.1) | 1.32 (0.93, 1.88) | 1.60 (1.07, 2.38) |
| Receipt of Omicron-targeted vaccines<sup>3</sup> | 0 Omicron-targeted vaccine doses | 3,096 (67.1) | 1,927 (62.6) | ref. | ref. |
|  | Any Omicron-targeted vaccine | 1,518 (32.9) | 1,153 (37.4) | 1.13 (1.05, 1.22) | 1.12 (1.03, 1.21) |
|  | Both BA.4/BA.5 (bivalent) and XBB1.5 (monovalent) vaccines | 464 (10.1) | 432 (14.0) | 1.43 (1.15, 1.43) | 1.28 (1.13, 1.45) |
|  | No BA.4/BA.5 (bivalent) vaccine doses | 3,205 (69.5) | 1,996 (64.8) | ref. | ref. |
|  | Any BA.4/BA.5 (bivalent) vaccine doses | 1,409 (30.5) | 1,084 (35.2) | 1.14 (1.06, 1.23) | 1.10 (1.01, 1.20) |
|  | No XBB.1.5 (monovalent) vaccine doses | 4,041 (87.6) | 2,579 (83.7) | ref. | ref. |
|  | Any XBB.1.5 (monovalent) vaccine doses | 573 (12.4) | 501 (16.3) | 1.23 (1.11, 1.35) | 1.14 (1.01, 1.28) |
| Timing of prior vaccination<sup>3</sup> | No doses received | 569 (12.3) | 272 (8.8) | ref. | ref. |
|  | Last vaccine dose within <3 months | 527 (11.4) | 436 (14.2) | 1.18 (1.07, 1.31) | 1.02 (0.79, 1.30) |
|  | Last vaccine dose within 3-6 months | 79 (1.7) | 92 (3.0) | 1.35 (1.10, 1.66) | 1.07 (0.85, 1.34) |
|  | Last vaccine dose >6 months prior | 3,439 (74.5) | 2,280 (74.0) | 0.98 (0.90, 1.06) | 0.86 (0.61, 1.22) |
| Documented prior infection | 0 documented infections | 2,505 (54.3) | 1,506 (48.9) | ref. | ref. |
|  | Any prior infection | 2,109 (45.7) | 1,574 (51.1) | 1.14 (1.06, 1.22) | 1.09 (1.02, 1.18) |
|  | 1 documented infection | 1,753 (38.0) | 1,269 (41.2) | 1.11 (1.03, 1.20) | 1.08 (1.00, 1.17) |
|  | 2 documented infections | 332 (7.2) | 278 (9.0) | 1.17 (1.03, 1.33) | 1.13 (0.99, 1.29) |
|  | ≥3 documented infections | 24 (0.5) | 27 (0.9) | 1.37 (0.93, 2.00) | 1.30 (0.89, 1.91) |

Data encompass the primary analytic cohort, comprised of individuals testing positive for SARS-CoV-2 from tests undertaken in outpatient settings between 1 December, 2023 and 30 January, 2024 which were processed via TaqPath COVID-19 Combo Kit assays, who belonged to KPSC health plans for at least one year prior to their index test date.
<sup>1</sup>Unadjusted odds ratios are computed via conditional logistic regression models matching on week of testing alone.
<sup>2</sup>Adjusted odds ratios are computed via conditional logistic regression models matching on week of testing and controlling for age, sex, race/ethnicity, body mass index, history of cigarette smoking, prior-year healthcare utilization across all settings, Charlson comorbidity index, and median household income within cases' census tract according to the categorization scheme indicated in **Table 1**. Missing values were addressed via multiple imputation, with results pooled across 5 pseudo-dataset replicates.
<sup>3</sup>Analyses of vaccine type and timing adjust for number of monovalent wild-type (Wuhan-Hu-1) vaccine doses received.

When distinguishing vaccines by type, cases infected with BA.2.86-derived lineages had 12% (3-21%) higher adjusted odds of having received any Omicron-targeted vaccine doses than cases infected with non-BA.2.86 lineages (**Table 2**). Adjusted odds of having received both BA.4/BA.5-targeted bivalent and XBB.1.5-targeted monovalent vaccine doses (versus no Omicron-targeted vaccine doses) were 28% (13-45%) higher among cases infected with BA.2.86-derived lineages than among cases infected with non-BA.2.86 lineages; cases infected with BA.2.86-derived lineages also had 10-14% higher odds of having received each of these vaccines independently in comparison to cases infected with non-BA.2.86 lineages. Following adjustment for the number and type of vaccine doses received, differences in timing of cases’ most recent COVID-19 vaccine doses were not apparent between cases infected with BA.2.86-derived lineages or non-BA.2.86 lineages.

We also compared history of documented prior SARS-CoV-2 infection among cases infected with BA.2.86-derived lineages and non-BA.2.86 lineages, recognizing that under-detection of such infections could lead to bias in estimates of effect sizes. Among cases infected with BA.2.86-derived lineages and non-BA.2.86 lineages, 54% and 49%, respectively, had no documented history of SARS-CoV-2 infection (**Table 2**); adjusted odds of any documented prior infection were 9% (2-18%) higher among cases infected with BA.2.86-derived lineages than among cases infected with non-BA.2.86 lineages. Point estimates of the association of infecting lineage with cases’ number of documented prior infections were consistent with a dose-response relationship, although statistical precision was limited. Adjusted odds of 1, 2, and ≥3 prior documented infections were 8% (0-17%), 13% (−1-29%) and 30% (−11-91%) higher among cases infected with BA.2.86-derived lineages than among cases infected with non-BA.2.86-derived lineages.

In analyses distinguishing the periods during which cases’ prior infections occurred, documented infection during the period when XBB lineages were dominant in circulation (1 December, 2022 to 31 October, 2023) was more common among cases infected with BA.2.86-derived lineages than among cases infected with non-BA.2.86 lineages (adjusted odds ratio = 1.16 [1.02-1.32]; **Table S4**). However, documented prior infection during the period when BA.2 lineages were dominant (3 February to 24 June, 2022) was also more common among cases infected with BA.2.86-derived lineages than among cases infected with non-BA.2.86 lineages (adjusted odds ratio = 1.16 [1.02-1.32]), suggesting that responses to infection with ancestral BA.2 lineages did not confer greater protection against BA.2.86-derived lineages in comparison to co-circulating XBB-derived lineages.

### Comparison of immune history by calendar period

To overcome limitations in statistical power affecting comparisons within the primary analytic cohort, we also assessed outcomes among cases diagnosed in all outpatient settings over bimonthly intervals throughout the study period. Although determination of individual-level lineage was not possible for cases tested on assays without SGTF results, we expected that differences in immune history among cases infected with BA.2.86-derived lineages and non-BA.2.86 lineages would be reflected among cases diagnosed at differing points in time during expansion of the JN.1 lineage. Consistent with our primary results obtained at the level of individual infection genotype, cases diagnosed at later points in time (i.e., as BA.2.86-derived lineages became dominant in circulation) tended to have received greater numbers of COVID-19 vaccine doses, and to have experienced greater numbers of prior documented SARS-CoV-2 infections (through the period ending 31 October, 2023) in comparison to cases diagnosed in November, 2023 (**Figure 2**). Compared to those diagnosed in November, 2023, cases diagnosed between 16-30 January, 2024 had 18% (4-34%) and 39% (14-70%) higher odds of having received 6 and ≥7 COVID-19 vaccine doses, respectively. Similarly, adjusted odds of 1, 2, and ≥3 prior documented SARS-CoV-2 infections were 13% (8-19%), 18% (7-29%), and 28% (–1-67%) higher among cases diagnosed between 16-30 January, 2024 in comparison to those diagnosed between 1-30 November, 2023. Implementation of XBB-targeted monovalent COVID-19 vaccines throughout Fall, 2023 prevented similar period-based comparisons for receipt of updated vaccines.

**Figure 2:**
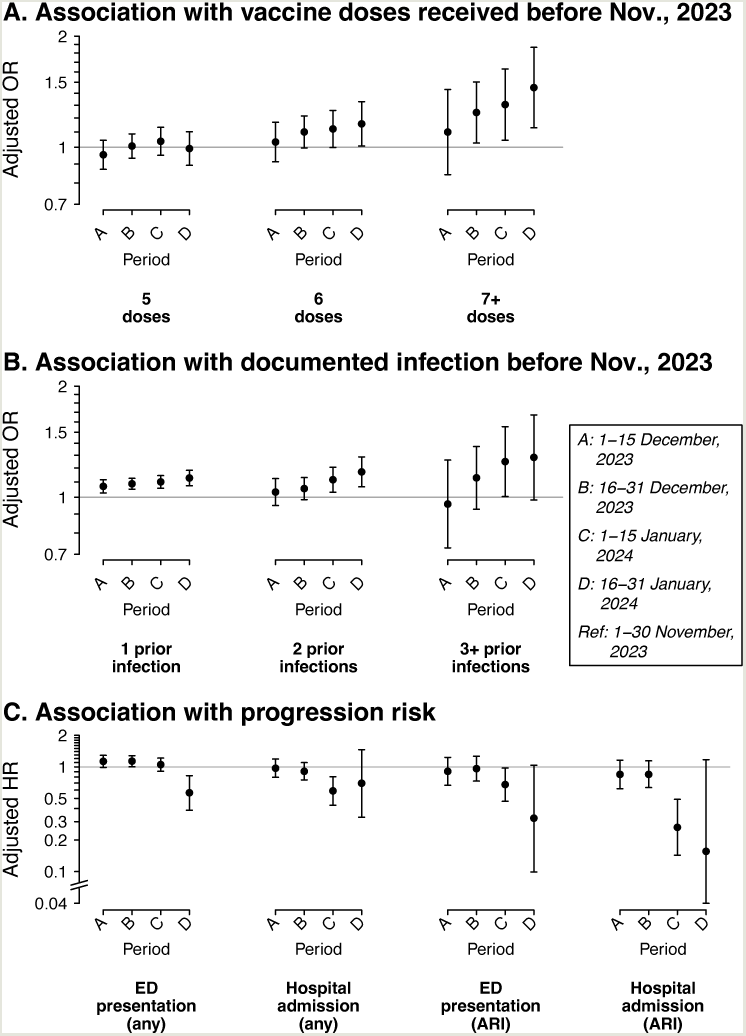
Period-based comparison of prior vaccination, prior documented infection, and risk of progression to various clinical outcomes. Panels illustrate (**a**) adjusted odds ratios, fitted via logistic regression models, for receipt of 5, 6, or ≥7 COVID-19 vaccine doses (relative to zero doses) among all outpatient cases diagnosed in the indicated periods relative to those diagnosed between 1-30 November, 2023; (**b**) adjusted odds ratios, fitted via logistic regression models, for documentation of 1, 2, or ≥3 prior SARS-CoV-2 infections (relative to zero documented prior SARS-CoV-2 infections) among all outpatient cases diagnosed in the indicated periods relative to those diagnosed between 1-30 November, 2023; and (**c**) adjusted hazard ratios, fitted via Cox proportional hazards models, for progression to emergency department (ED) presentation or hospital admission, due to any cause or in association with acute respiratory infection (ARI) diagnoses, comparing outpatient cases diagnosed in the indicated periods to those diagnosed between 1-30 November, 2023. All models adjust for age, sex, race/ethnicity, body mass index, history of cigarette smoking, prior-year healthcare utilization across all settings, Charlson comorbidity index, and median household income within cases’ census tract according to the categorization scheme indicated in **Table 1**. In addition, Cox proportional hazards models adjust for nirmatrelvir-ritonavir receipt as a time-varying exposure. Missing values were addressed via multiple imputation, with results pooled across 5 pseudo-dataset replicates.

### Comparison of clinical outcomes by infecting lineage

We next assessed risk of clinical outcomes signifying disease progression following an initial outpatient diagnosis within our primary analytic cohort. Outcomes of interest included emergency department presentation within 14 days, hospital admission within 28 days, and intensive care unit (ICU) admission, mechanical ventilation, or death within 60 days. Rates of these outcomes were 24.6, 4.0, and 0.8 events, respectively, per 10,000 person-days of follow-up among cases infected with non-BA.2.86 lineages and 11.4, 1.6, and 0.4 events, respectively, per 10,000 person-days among cases infected with BA.2.86-derived lineages (**Table 3**). In Cox proportional hazards models matching cases on testing week and controlling for measured case characteristics including vaccination and documented prior infection (see **Methods**), adjusted hazards of emergency department presentation and hospital admission were 54% (32-69%) and 51% (–15-79%) lower, respectively, among cases infected with BA.2.86-derived lineages than among cases infected with non-BA.2.86 lineages. Adjustment for risk factors was not feasible for analyses of ICU admission, mechanical ventilation, or death due to the low frequency of such outcomes. Expecting that some emergency department presentations and hospital admissions following outpatient SARS-CoV-2 detections would be attributable to factors unrelated to COVID-19, we also conducted analyses restricting outcomes to emergency department presentations or hospital admissions associated with acute respiratory infection (ARI) diagnosis codes (**Table S5**). Adjusted hazards of ARI-associated emergency department presentations and hospital admissions were 62% (–2-86%) and 85% (–12-98%) lower, respectively, among cases infected with BA.2.86-derived lineages than among cases infected with non-BA.2.86 lineages.

**Table 3:** Clinical progression among individuals infected with SARS-CoV-2 according to infecting lineage.

| Episode type | Outcome | Events, *n* (Rate per 10,000 days) |  | Hazard ratio (95% CI), JN.1 vs. non-JN.1 infection |  |
| --- | --- | --- | --- | --- | --- |
|  |  | S-gene detected<br>(non-BA.2.86 lineage)<br>*N*=4,614 | S-gene target failure<br>(BA.2.86-derived lineage)<br>*N*=3,080 | Unadjusted<sup>1</sup> | Adjusted<sup>2</sup> |
| Episodes associated with all causes | Emergency department presentation | 126 (24.6) | 33 (11.4) | 0.41 (0.27, 0.60) | 0.46 (0.31, 0.68) |
|  | Hospital admission | 36 (4.0) | 7 (1.6) | 0.33 (0.15, 0.76) | 0.49 (0.21, 1.15) |
|  | ICU admission, mechanical ventilation, or death | 9 (0.8) | 2 (0.4) | 0.40 (0.08, 1.89) | — |
|  | Death | 5 (0.5) | 1 (0.2) | 0.38 (0.04, 3.33) | — |
| ARI-associated episodes<sup>3</sup> | Emergency department presentation | 22 (4.2) | 5 (1.7) | 0.35 (0.13, 0.96) | 0.38 (0.14, 1.02) |
|  | Hospital admission | 18 (2.0) | 1 (0.2) | 0.09 (0.01, 0.68) | 0.15 (0.02, 1.12) |

Data encompass the primary analytic cohort, comprised of individuals testing positive for SARS-CoV-2 from tests undertaken in outpatient settings between 1 December, 2023 and 30 January, 2024 which were processed via TaqPath COVID-19 Combo Kit assays, who belonged to KPSC health plans for at least one year prior to their index test date.
<sup>1</sup>Unadjusted hazard ratios are computed via Cox proportional hazards regression models matching on week of testing alone.
<sup>2</sup>Adjusted odds ratios are computed via Cox proportional hazards regression models matching on week of testing and controlling for age, sex, race/ethnicity, body mass index, history of cigarette smoking, prior-year healthcare utilization across all settings, Charlson comorbidity index, and median household income within cases' census tract according to the categorization scheme indicated in **Table 1**. In addition, nirmatrelvir-ritonavir receipt is defined as a time-varying exposure. Missing values were addressed via multiple imputation, with results pooled across 5 pseudo-dataset replicates.
<sup>3</sup>Acute respiratory infection diagnosis codes are presented in **Table S5**.

### Comparison of clinical outcomes by calendar period

While the above findings were consistent with a scenario in which BA.2.86-derived lineages were associated with attenuated clinical severity, comparisons for most outcomes lacked sufficient statistical power to exclude the possibility of no difference. To overcome this limitation, we next compared outcomes among cases diagnosed during successive bimonthly periods throughout the study period as JN.1 became the dominant circulating variant (**Figure 2**). Compared to cases tested between 1-30 November, 2023, adjusted hazard ratios for emergency department presentation were 1.05 (0.91-1.21) for cases tested between 1-15 January, 2024 and 0.57 (0.39-0.82) for cases tested between 16-30 January, 2024. For the outcome of ARI-associated emergency department presentations, adjusted hazards ratios declined to 0.68 (0.47-0.97) and 0.32 (0.10-1.04), respectively, for cases tested between 1-15 January and 16-30 January, 2024. For the same periods, adjusted hazard ratios of hospital admission were 0.59 (0.43-0.80) and 0.70 (0.33-1.46), respectively, and adjusted hazard ratios of ARI-associated hospital admission were 0.26 (0.14-0.49) and 0.16 (0.05-1.17), respectively.

In interpreting findings of the period-based analysis, it is important to consider that differences over time in the clinical threshold at which cases sought SARS-CoV-2 testing or subsequently presented for care—especially during the holiday season (20)—could hinder attribution of differences in risk by calendar period to emergence of the JN.1 lineage. For instance, we observed transient increases in risk of emergency department presentation and hospital admission or ARI-associated hospital admission among cases diagnosed between December 7-10 and December 24-31, 2023 during the Hannukah and Christmas/New Year holidays, respectively (**Figure 3**); this outcome may thus owe to seasonal behavioral factors, such as seeking care only in the context of relatively severe illness during the holiday season.

**Figure 3:**
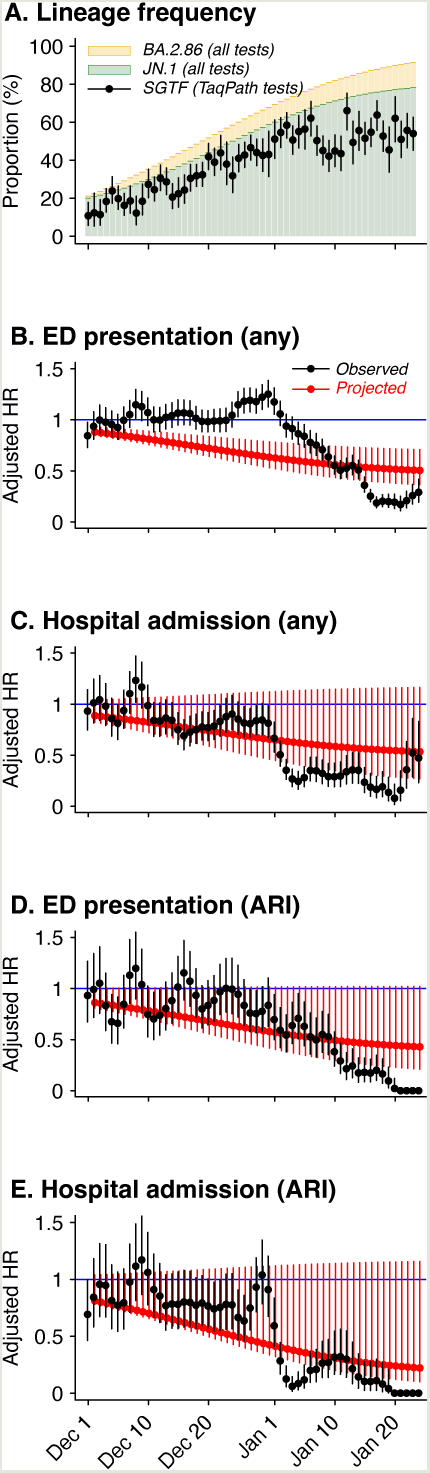
Observed and projected changes in risk of progression. Panels illustrate (**a**) the daily proportion of outpatient tests exhibiting S-gene target failure (BA.2.86-derived lineages) among all tested within the primary analytic cohort; (**b**) estimates of the observed day-specific adjusted hazard ratio of emergency department (ED) presentation due to any cause (black), as well as projected estimates of the day-specific adjusted hazard ratio of ED presentation due to any cause resulting only from changes in lineage composition among outpatient-diagnosed cases (defined as 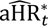 in the **Methods**; red); (**c**) for the outcome of hospital admission due to any cause, corresponding estimates of the observed day-specific hazard ratios and projected adjusted hazard ratios based only on changes in lineage composition (black and red, respectively); (**d**) for the outcome of ED presentations associated with acute respiratory infection (ARI) diagnoses, corresponding estimates of the observed day-specific hazard ratios and projected adjusted hazard ratios based only on changes in lineage composition (black and red, respectively); and (**e**) for the outcome of hospital admissions associated with acute respiratory infection (ARI) diagnoses, corresponding estimates of the observed day-specific hazard ratios and projected adjusted hazard ratios based only on changes in lineage composition (black and red, respectively). For all panels, points and lines denote median estimates and accompanying 95% confidence intervals, respectively.

To understand the potential magnitude of changes in risk attributable to lineage replacement, we compared adjusted hazard ratios of each clinical outcome among all outpatient-diagnosed cases (estimated daily relative to risk among cases diagnosed between 1-30 November, 2023) to a projected adjusted hazard ratio for the same day under a scenario without changes over time in healthcare-seeking behavior. We obtained this counterfactual risk by weighting estimates of the adjusted hazard ratio for each outcome associated with BA.2.86-derived lineages by the daily proportion of cases found to be infected with BA.2.86-derived lineages among those randomly selected for sequencing (see **Methods**). Using this approach, we projected that changes in lineage composition alone would cause cases diagnosed 15 January, 2024 to experience 47% (27-60%) and 54% (−2-74%) lower risk of emergency department presentation associated with any cause and with ARI, respectively, in comparison to cases diagnosed between 1-30 November, 2023. In contrast, we observed 64% (56-71%) and 82% (68-91%) lower risk of any and ARI-associated emergency department presentation, respectively, among cases diagnosed 15 January, 2024 compared to those diagnosed between 1-30 November, 2023. Similarly, projected and observed reductions in risk were 44% (−15-69%) and 76% (62-87%), respectively, for any hospital admission, and 73% (–15-85%) and 90% (72-98%), respectively, for ARI-associated hospital admission. Thus, reductions in risk of severe outcomes over the study period could be explained only partially by expansion of BA.2.86-derived lineages.

### Sensitivity analyses addressing protection against severe disease due to unobserved prior infections

Our inability to control completely for cases’ infection history further limits our comparison of clinical outcomes according to infecting lineage. In the event that cases infected with BA.2.86 experienced greater numbers of unobserved as well as observed infections in comparison to cases infected with non-BA.2.86 lineages, protection from these unobserved infections could contribute to the apparent association of infecting lineage (or infection during BA.2.86-dominant periods) with attenuated risk of clinical progression (21). Restricting the sample to cases who had >5 healthcare interactions in the preceding year (*N*=5,167 members of the primary analytic cohort)—among whom we expected that prior infections would have been recorded with greater likelihood—yielded results confirming those of the primary analyses (**Table S6**), although this approach was not guaranteed to eliminate potential bias due to unrecorded infections. We therefore undertook sensitivity analyses imputing alternative individual-level infection histories to account for the possibility that unobserved prior infections were especially prevalent among individuals infected with BA.2.86-derived lineages and those who evaded severe outcomes (see **Methods**).

Extreme conditions of differential ascertainment of prior infections were needed to arrive at scenarios in which BA.2.86- derived lineages were not associated with attenuated risk of clinical progression. For both outcomes, the direction of association was reversed (resulting in a statistically significant association of BA.2.86-derived lineages with increased risk of disease progression) only in contexts where the true number of prior infections among cases infected with BA.2.86-derived lineages who avoided the need for emergency department or inpatient care was 6-27 times higher than that observed (**Figure 4**). Under such conditions, the mean number of prior infections among cases infected with BA.2.86- derived lineages spanned 6.6-16.7, and exceeded the mean number of prior infections among cases infected with non-BA.2.86 lineages by a factor of 3.5-4.2, or 5.0-11.8 total unascertained infections. Cases infected with BA.2.86-derived lineages who avoided emergency department presentation or hospital admission would have experienced 5.6-11.5 more infections, on average, than those who experienced these outcomes.

**Figure 4:**
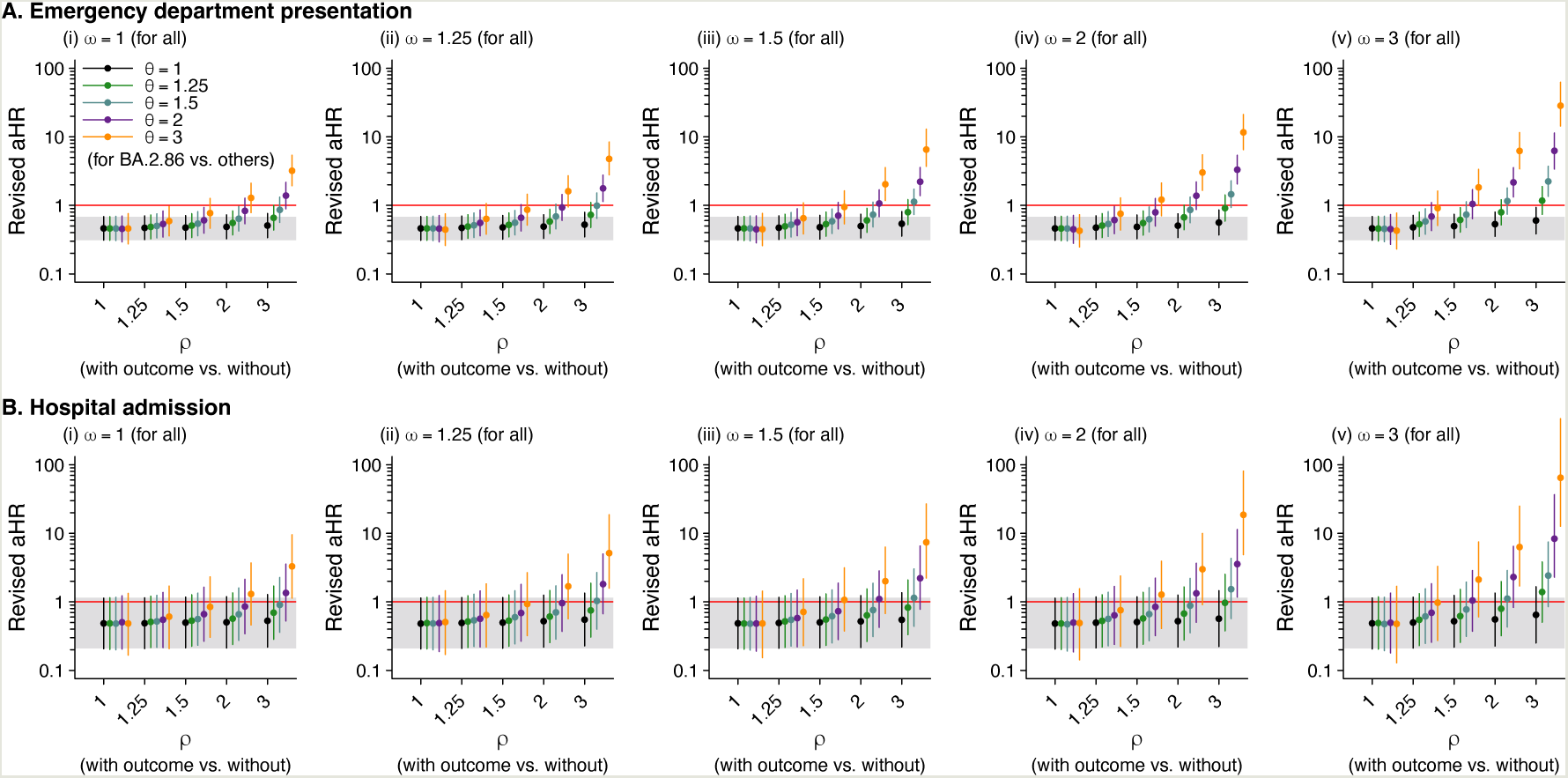
Sensitivity analyses addressing the association of SGTF with risk of emergency department presentation and hospital admission in the presence of differential misclassification of prior infection according to infecting lineage and clinical outcome. We illustrate estimates of the adjusted hazard ratio of progression to (**a**) emergency department presentation and (**b**) hospital admission under analyses imputing “full” infection histories for cases under the assumption of differential misclassification of prior infection status. We consider multipliers of 1, 1.25, 1.5, 2, and 3 for the ratio of true to observed infections, first non-differentially among all cases (𝜔), and for the relative ratio of true to observed infections comparing cases who evaded the indicated outcome versus those who experienced it (𝜌) and comparing cases infected with BA.2.86-derived lineages to those infected with non-BA.2.86 lineages (𝜃). Points and lines illustrate median estimates with accompanying 95% confidence intervals. Grey bands illustrate 95% confidence intervals for estimates from the primary analysis.

While cases’ true number of unascertained prior infections cannot be known, these figures likely exceed plausible levels based on estimates of SARS-CoV-2 reporting completeness during various phases of the pandemic (22,23). Among all cases in the study population, only 0.6%, 0.06%, and 0.0005% were observed to have experienced 3, 4, or 5 prior infections throughout follow-up. Furthermore, mean numbers of documented prior infections were only 1.2-fold higher among cases infected with BA.2.86-derived lineages than among cases infected with non-BA.2.86 lineages (0.6 and 0.5 documented prior infections on average, respectively, in the two case populations).

## DISCUSSION

Our study identifies escape of immune responses derived from prior COVID-19 vaccination or SARS-CoV-2 infection as a likely factor in the emergence of BA.2.86/JN.1 lineage during the period from December, 2023 to January, 2024. Cases infected with BA.2.86-derived lineages had 38%, 51%, and 60% higher adjusted odds of having received 5, 6, and ≥7 COVID-19 vaccine doses in comparison to cases infected with co-circulating lineages, predominantly descending from XBB. Although under-detection of prior infections limited our ability to compare infection history among cases infected with BA.2.86-derived lineages or non-BA.2.86 lineages, cases infected with BA.2.86-derived lineages had at least 8%, 13%, and 30% higher adjusted odds of having experienced 1, 2, or ≥3 prior documented SARS-CoV-2 infections. Overcoming concerns about statistical power within these primary analyses, our findings were reflected in period-based analyses in which prior vaccination and infection were each more strongly associated with diagnosis during phases when the JN.1 lineage accounted for a greater share of new SARS-CoV-2 infections. These findings suggest that immune responses resulting from prior vaccination or infection may have conferred greater protection against infection with XBB-derived lineages than BA.2.86-derived lineages.

Our findings that cases infected with BA.2.86-derived lineages had greater odds of prior infection during periods when XBB lineages were dominant in circulation, and greater odds of having received Omicron-targeted COVID-19 vaccine doses, are consistent with previous evidence of immune evasion by BA.2.86 and the JN.1 lineage, in particular. Sera from individuals previously infected with XBB.1.5 lineages have showed greater capacity to neutralize EG.5 and other XBB “FLip” lineages in comparison to BA.2.86 (24). Moreover, sera from XBB.1.5-targeted monovalent vaccine recipients exhibited superior neutralization of the XBB-derived EG5.1 and HK.3 lineages in comparison to sera from BA.4/BA.5- targeted bivalent vaccine recipients (25); in contrast, sera from recipients of XBB.1.5-targeted and BA.4./BA.5-targeted vaccines had weak, non-differential capacity to neutralize the JN.1 lineage. On the surface, greater frequency of prior infection during the XBB-dominant period among cases infected with BA.2.86-derived lineages appears consistent with a scenario in which ancestral XBB lineages induced specific cross-protection against descendant lineages such as EG.5, HK.3, HV.1, JD.1, and JG.3. However, we also identified that cases infected with BA.2.86-derived lineages were more likely to have been infected during the BA.2-dominant period. Thus, differences in the ability of BA.2.86-derived lineages and non-BA.2.86 lineages to evade immune responses associated with prior XBB infection may be comparable to differences in their ability to evade immune responses associated with ancestral BA.2 lineages. Unfortunately, changes in testing effort over time impede direct comparison of effect size estimates for associations of infecting lineage with documented infection during differing periods of the COVID-19 pandemic.

In an early study including 679 COVID-19 cases, XBB.1.5-targeted vaccination showed modestly weaker effectiveness against infection with BA.2.86-derived lineages in comparison to XBB-derived lineages (49% vs. 60%; (19)), consistent with findings from a cohort study in which recipients of XBB.1.5-targeted monovalent vaccine had lower odds of infection with XBB-derived lineages in comparison to BA.2.86-derived lineages (26). In a larger study of 6,551 adults with clinically-attended COVID-19, XBB.1.5-targeted monovalent vaccine conferred weaker effectiveness against progression from outpatient to emergency department or inpatient levels of care delivery among cases infected with BA.2.86-derived lineages than among cases infected with XBB-derived lineages (38% vs. 72%; (27)). Individuals with a history of XBB infection, along with recipients of BA.4/BA.5-targeted or XBB.1.5-targeted vaccines, may thus have provided a niche facilitating the expansion of BA.2.86-derived lineages including JN.1 over existing XBB-derived strains.

Several findings from our study suggested attenuation of disease severity in BA.2.86-derived lineages. First, we estimated that cases diagnosed in outpatient settings who were infected with BA.2.86-derived lineages had 54% lower risk of emergency department presentation than cases who were infected with non-BA.2.86 lineages. We obtained similar point estimates of differences in risk of progression to hospital admission, and greater point estimates of differences in risk for outcomes associated with ARI diagnoses. However, statistical power was constrained by the limited number of cases experiencing progression to these outcomes. Period-based analyses revealed continuous reductions in cases’ risk of ARI-associated emergency department presentation and hospital admission throughout the period of JN.1 expansion, although changes over time in cases’ risk of progression exceeded expectations based on lineage replacement alone. This may reflect differences over time in the clinical threshold at which cases sought testing, or other unmeasured differences over time in the characteristics of cases becoming infected; alternatively, the distribution of BA.2.86-derived sublineages identified among the primary analytic cohort may have differed from the distribution within the full case population, particularly by the end of the study period. While our inability to accurately classify cases’ infection history may limit interpretation of our findings, sensitivity analyses identified that implausible numbers of unobserved infections would be needed among cases infected with BA.2.86-derived lineages to reverse the direction of observed associations between infecting lineage and disease progression (mean 6.6-16.7 prior infections). No epidemiologic evidence supports a scenario in which appreciable numbers of individuals would have experienced this many SARS-CoV-2 infections prior to the study period. Further, the lack of meaningful differences in demographics, clinical characteristics, or healthcare-seeking behavior prior to diagnosis among cases infected with BA.2.86-derived lineages and non-BA.2.86 lineages makes it unlikely that the proportion of prior infections that were ascertained should differ so appreciably between these groups.

Several additional limitations should be considered. First, as our study period coincided with multiple holidays, clinical thresholds at which individuals sought SARS-CoV-2 testing and presented for subsequent care may have varied over time. While this poses limitations for period-based analyses, matching cases on their week of testing is likely to have alleviated resulting bias in primary analyses. Moreover, KPSC maintained consistent criteria for hospital admission throughout the study period, and did not experience surges in severe COVID-19 cases that would necessitate tightening of such criteria. Second, our sample was identified through outpatient clinical testing, and may represent a more severe spectrum of all infections than what we would expect to identify through active, prospective testing of asymptomatic as well as symptomatic individuals. Thus, rates of progression to each clinical outcome should not be generalized to all infections. Third, because only a small proportion of samples were submitted for sequencing, we cannot distinguish clinical outcomes and characteristics of cases infected with JN.1 or other BA.2.86-derived lineages. Fourth, restricting our study to individuals with confirmed SARS-CoV-2 infection represents an instance of conditioning on a post-exposure variable for analyses of vaccination or prior infection (28). Regardless, our case-only approach offered the advantage of selecting on healthcare-seeking behavior among cases regardless of their infecting lineage, which may otherwise represent a key source of bias in comparisons of individuals with differing histories of vaccination and prior infection (29). Relatedly, our study framework comparing is observational in nature, and may be subject to unmeasured sources of confounding.

Risk of adverse clinical outcomes among cases diagnosed with SARS-CoV-2 infection in outpatient settings was low in our study population relative to those infected during earlier phases of the COVID-19 pandemic. Despite evidence from our study that BA.2.86-derived lineages may partially evade immunity acquired through prior vaccination (including with XBB.1.5-targeted boosters) and infection, and external evidence of extensive community transmission of these lineages (8), rates of COVID-19 hospital admissions and mortality did not match burden experienced during expansion of the Omicron BA.1, BA.4/BA.5, and XBB.1.5 lineages. Continued monitoring of risk of these clinical outcomes as well as vaccine effectiveness remains important to inform response strategies to novel SARS-CoV-2 lineages, including the need for reformulation of booster doses.

## METHODS

### Study population

Our study included all individuals who received positive molecular tests for SARS-CoV-2 infection between 1 November, 2023 and 30 January, 2024, who had no clinical record of receiving any positive SARS-CoV-2 test or COVID-19 diagnosis within 90 days before their first test (“index test”) within this period, and who were enrolled in KPSC health plans for ≥1 year before their index test (allowing for lapses in membership of up to 45 days). We restricted the primary analytic cohort to individuals tested between 1 December, 2023 and 30 January, 2024 whose index tests were processed using ThermoFisher TaqPath COVID-19 Combo Kit assays, for whom SGTF readout was available. Cases belonging to the primary analytic cohort were predominantly tested in outpatient-serving facilities without in-house laboratory facilities, which relied on regional centers using the ThermoFisher TaqPath COVID-19 Combo Kit for specimen processing. Cases tested in hospital-based emergency departments had specimens processed in hospital laboratories via devices without SGTF readout.

### Exposures

We defined prior vaccine doses as those received >14 days before individuals’ index test and distinguished doses received as monovalent wild-type (Wuhan-Hu-1) vaccines, bivalent BA.4/BA.5/Wuhan-Hu-1 vaccines, and monovalent XBB.1.5 vaccines. We defined prior infections as laboratory-confirmed SARS-CoV-2 diagnoses without any positive SARS-CoV-2 test result or COVID-19 diagnosis within the preceding 90 days. We accounted for nirmatrelvir-ritonavir receipt as a time-varying exposure for all dispenses initiated within 5 days after the index test date; for dispenses initiated after the index date, individuals’ exposure status was permitted to change beginning on the dispense date. Additional characteristics obtained from patient electronic health records and accompanying demographic metadata included cases’ age (categorized in 10-year increments for all analyses), sex, race/ethnicity (categorized as White non-Hispanic, Black non-Hispanic, Hispanic of any race, Asian, Pacific Islander, or other/mixed/unknown race/ethnicity), body mass index (categorized as underweight, normal weight, overweight, or obese if measured in the preceding year), history of cigarette smoking (current, former, or never smokers), prior-year healthcare utilization (categorized across outpatient, emergency department, and inpatient settings as presented in **Table 1**), Charlson comorbidity index (0, 1-2, 3-5, or ≥6), and median household income within their census tract (categorized as presented in **Table 1**).

### Outcomes

Within the primary analytic cohort, we considered cases with positive detection (c*T*<37) of the SARS-CoV-2 S, N, and orf1a/b genes to be infected with non-BA.2.86 lineages. We considered cases with positive detection of N and orf1a/b genes but no detection (c*T*≥37) of the S gene (SGTF) to be infected with BA.2.86-derived lineages. We followed cases for the following outcomes within the specified time range from the index test: any emergency department presentation within 14 days; ARI-associated emergency department presentation within 14 days; any hospital admission within 28 days; ARI-associated hospital admission within 28 days; ICU admission within 60 days; initiation of mechanical ventilation within 60 days; and death within 60 days. Due to the low frequency of severe outcomes, we defined a composite outcome of ICU admission, initiation of mechanical ventilation, or death within 60 days. For each case, we defined analysis periods ending at occurrence of any study outcome or censoring due to disenrollment; a new observation window was initiated at the point of nirmatrelvir-ritonavir dispense if treatment preceded any study outcome. For analyses of ARI-associated emergency department presentations and hospital admissions, we also censored observations at occurrence of the outcome without any accompanying ARI diagnosis code.

### Missing data

Among 68,281 cases, 14,563 (21%) had missing entries for at least one analytic variable. Missing value frequencies were as follows: 9,630 (14%) for body mass index, 8,535 (12.5%) for cigarette smoking, 4,129 (6%) for census tract median household income, and 1,102 (2%) for age. We populated 5 complete pseudo-datasets via multiple imputation using the Amelia package (30). For all analyses, we pooled results from replications across each pseudo-dataset.

### Comparison of prior vaccination and infection

For members of the primary analytic cohort, we fit conditional logistic regression models defining infection with BA.2.86-derived lineages or non-BA.2.86 lineages as the outcome variable and defining matching strata for their week of testing. Models controlled for age, sex, race/ethnicity, body mass index, history of cigarette smoking, prior-year healthcare utilization across all settings, Charlson comorbidity index, and median household income within cases’ census tract. Individuals were excluded from analyses if they received any vaccine dose within ≤14 days of their index test.

Primary analyses included count variables for doses of all COVID-19 vaccines received. We also report results of analyses which distinguished counts of monovalent wild-type (Wuhan-Hu-1) vaccine doses, BA.4/BA.5-targeted bivalent vaccine doses, and XBB.1.5-targeted vaccine doses, as well as the timing of receipt of the most recent vaccine dose (<3 months, 3-6 months, or >6 months). Analyses distinguishing prior infection by periods when distinct SARS-CoV-2 variants were dominant in circulation (**Table S4**) defined no documentation of prior SARS-CoV-2 infection as the reference exposure.

For period-based analyses including all outpatient-diagnosed cases without restriction on test assay, we fit separate logistic regression models defining infection during each of the periods of 1-15 December, 2023, 16-31 December, 2023, 1-15 January, 2024, or 16-30 January, 2024 as outcomes (“1”), with infection during the period of 1-30 November, 2023 defined as the control outcome (“0”). Models again included the number of vaccine doses cases had received and the number of recorded infections preceding cases’ index test. Cases who received vaccination on or after 1 November, 2023 were excluded to enable comparisons of exposures that all cases were eligible to encounter. Consistent with primary analyses, models controlled for age, sex, race/ethnicity, body mass index, history of cigarette smoking, prior-year healthcare utilization across all settings, Charlson comorbidity index, and median household income within cases’ census tract. No adjustment for calendar time was included due to our specification of infection timing as the outcome variable.

### Comparison of clinical outcomes

Within the primary analytic cohort, we fit Cox proportional hazards models for each outcome defining matching strata on cases’ week of testing. We defined infection with BA.2.86-derived lineages or non-BA.2.86 lineages as the exposure of interest. Models adjusted for the same covariates listed above for conditional logistic regression analyses, with the addition of a time-varying covariate for receipt of nirmatrelvir-ritonavir.

For period-based analyses including outpatient-diagnosed cases without restriction on test assay, we defined infection during the periods of 1-30 November, 2023 (reference period), 1-15 December, 2023, 16-31 December, 2023, 1-15 January, 2024, or 16-30 January, 2024 as the exposures of interest. We controlled for the same covariates as those included in primary analyses, with no adjustment for calendar time. Again, cases were excluded if they received vaccination after 1 November, 2023.

To distinguish the role of both secular (time-varying) factors and infecting lineage in contributing to the reduced incidence of severe disease outcomes over the course of the study period, we also compared empirical estimates of day-specific hazard ratios for cases’ risk of progression to each outcome to projections of day-specific hazard ratios based only on changes in SARS-CoV-2 lineage composition. Empirical estimates were fitted via Cox proportional hazards that included day-specific intercepts for each day from 1 December, 2023 through 30 January, 2024 (measured relative to risk for cases diagnosed between 1-30 November, 2023), and controlled for the same factors listed above. We projected corresponding day-specific estimates of the hazard ratio of progression due only to changes in SARS-CoV-2 lineage composition, 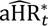, via the formula

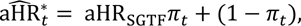

where aHR_SGTF_ indicated the adjusted hazard ratio of progression to the outcome of interest comparing cases infected with BA.2.86-derived lineages to cases infected with non-BA.2.86 lineages (as estimated in the primary analytic cohort), and 𝜋_𝑡_ indicated the proportion of cases infected at time *t* with BA.2.86-derived lineages among all cases for whom sequencing results were available. We generated day-specific estimates of 𝜋_𝑡_ by fitting a regression model to data representing weekly proportions of sequences found to represent BA.2.86-derived lineages; we defined polynomial transformations of calendar time as the independent variables, finding that a 5^th^-degree polynomial yielded the lowest value of the Bayesian information criterion (**Figure S1**). We overlay our projected 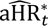 estimates with empirical day-specific adjusted hazard ratio estimates in **Figure 3**.

### Sensitivity analysis

To assess the sensitivity of our estimates to differential misclassification (undercounting) of prior infections, particularly among cases infected with BA.2.86-derived lineages and those who evaded severe clinical outcomes, we also conducted analyses imputing alternative infection histories among cases. Defining the prior number of infections for case *i* within the primary analytic cohort as a Poisson random variable with the underlying rate 𝜆*_i_*, we sampled alternative infection histories 𝑋*_i_* as Poisson random variables according to

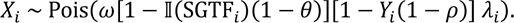

Here 𝜔 provided a multiplier conveying the minimum ratio of true to documented infections among all cases; if under-detection was considered differential for cases according to infecting lineage or a clinical outcome of interest, 𝜔 conveyed the ratio of true to documented infections among cases infected with non-BA.2.86 lineages (𝕀(SGTF*_i_*) = 0) who experienced the outcome (𝑌*_i_* = 1). The parameters 𝜃 and 𝜌, respectively, represented the relative ratio of true to documented infections among cases infected with BA.2.86-derived lineages (relative to cases infected with non-BA.2.86 lineages) and the relative ratio of true to documented infections among cases who evaded each clinical outcome (relative to those who experienced the outcome). We considered values of 1, 1.25, 1.5, 2, and 3 for 𝜔, 𝜌, and 𝜃, so that the corrected rate parameter could be up to 27 fold higher than that observed for cases infected with BA.2.86-derived lineages who evaded each clinical outcome.

We estimated 𝜆*_i_* via Poisson regression models defining cases’ number of prior infections as the outcome variable and including all other measured covariates as predictors. For each parameterization of {𝜔, 𝜌, 𝜃}, we drew 10 vectors of case infection histories *X̂_i_* and fit Cox proportional hazards models according to the specifications of the primary analysis.

### Software

We conducted analyses using R software (R Foundation for Statistical Computing, Vienna, Austria). We used the Amelia package (30) for multiple imputation and fit conditional logistic regression models and Cox proportional hazards models using the survival package (31).

## Data Availability

Individual-level testing and clinical outcomes data reported in this study are not publicly shared due to privacy protections for patient electronic health records. Individuals wishing to access disaggregated data, including data reported in this study, should submit requests for access to. Requests will receive a response within 14 days. De-identified data (including, as applicable, participant data and relevant data dictionaries) will be shared upon approval of analysis proposals with signed data-access agreements in place.
Analysis code is available from GitHub (https://github.com/joelewnard/jn1).

## ACKNOWLEDGMENTS

This study was funded by the US Centers for Disease Control and Prevention (SYT) and the US National Institutes of Health (R01-AI148336 to JAL). This study was reviewed by Kaiser Permanente Southern California and the US Centers for Disease Control and Prevention (CDC) and was conducted consistent with applicable federal law and CDC policy (45 CFR part 46, 21 CFR part 56; 42 USC Sect. 241 (d); 5 USC Sect. 552a; 44 USC Sect. 3501 et seq.).

## DISCLAIMER

The findings and conclusions in this report are those of the authors and do not necessarily represent the official position of the Centers for Disease Control and Prevention.

## RESOURCE SHARING

Individual-level testing and clinical outcomes data reported in this study are not publicly shared due to privacy protections for patient electronic health records. Individuals wishing to access disaggregated data, including data reported in this study, should submit requests for access to. Requests will receive a response within 14 days. De-identified data (including, as applicable, participant data and relevant data dictionaries) will be shared upon approval of analysis proposals with signed data-access agreements in place.

Analysis code is available from GitHub (https://github.com/joelewnard/jn1).

---

**Table S1:** Characteristics of outpatient cases tested on ThermoFisher TaqPath COVID-19 Combo Kit (TF) and other assays.

| Characteristic |  | Cases, n/N (%) |  |
| --- | --- | --- | --- |
|  |  | Primary analytic cohort (tested on TF assays) | Cases tested on non-TF assays |
|  |  | N=7,694 | N=38,373 |
| Age (years)<sup>1</sup> | 0-9 | 323 (4.2) | 1,056 (2.8) |
|  | 10-19 | 387 (5.0) | 1,406 (3.7) |
|  | 20-29 | 737 (9.6) | 2,995 (7.8) |
|  | 30-39 | 1,273 (16.5) | 5,679 (14.8) |
|  | 40-49 | 1,424 (18.5) | 6,502 (16.9) |
|  | 50-59 | 1,359 (17.7) | 6,726 (17.5) |
|  | 60-69 | 1,117 (14.5) | 6,408 (16.7) |
|  | 70-79 | 698 (9.1) | 4,883 (12.7) |
|  | ≥80 | 376 (4.9) | 2,718 (7.1) |
| Sex | Female | 4,648 (60.4) | 23,711 (61.8) |
|  | Male | 3,046 (39.6) | 14,662 (38.2) |
| Race | White, non-Hispanic | 1,669 (21.7) | 13,180 (34.3) |
|  | Black, non-Hispanic | 803 (10.4) | 2,607 (6.8) |
|  | Hispanic (any race) | 3,661 (47.6) | 15,371 (40.1) |
|  | Asian | 1,014 (13.2) | 4,976 (12.5) |
|  | Pacific Islander | 75 (1.0) | 294 (0.8) |
|  | Other/mixed/unknown race | 472 (5.5) | 2,125 (6.1) |
| Body mass index<sup>1</sup> | Underweight (<18.5) | 280 (3.6) | 1,204 (3.1) |
|  | Normal weight (18.5-24.9) | 1,583 (20.6) | 7,838 (20.4) |
|  | Overweight (25.0-29.9) | 2,077 (27.0) | 10,329 (26.9) |
|  | Obese (≥30.0) | 2,907 (37.8) | 14,533 (37.9) |
| Cigarette smoking<sup>1</sup> | Never smoker | 5,395 (70.1) | 25,622 (66.8) |
|  | Former smoker | 1,366 (17.8) | 7,530 (19.6) |
|  | Current smoker | 283 (3.7) | 1,354 (3.5) |
| Charlson comorbidity index | 0 | 4,740 (61.6) | 21,568 (56.2) |
|  | 1-2 | 2,098 (27.3) | 11,218 (29.2) |
|  | 3-5 | 616 (8.0) | 2,819 (10.0) |
|  | ≥6 | 240 (3.1) | 1,768 (4.6) |
| Prior-year healthcare utilization | 0-9 outpatient encounters | 3,979 (51.7) | 19,234 (50.1) |
|  | 10-19 outpatient encounters | 1,991 (25.9) | 10,352 (27.0) |
|  | 20-29 outpatient encounters | 893 (11.6) | 4,412 (11.5) |
|  | ≥30 outpatient encounters | 831 (10.8) | 4,375 (11.4) |
|  | Any emergency department presentation | 1,682 (21.9) | 9,002 (23.5) |
|  | Any inpatient admission | 418 (5.4) | 2,573 (6.7) |
| Census tract household median income<sup>1</sup> | <\$40,000 | 347 (4.5) | 1,421 (3.7) |
| | \$40,000-79,999 | 3,054 (39.7) | 13,379 (34.9) |
| | \$80,000-119,999 | 2,579 (33.5) | 13,555 (35.3) |
| | \$120,000-159,999 | 1,080 (14.0) | 6,345 (16.5) |
| | ≥\$160,000 | 361 (4.7) | 2,242 (5.8) |
| Receipt of nirmatrelvir-ritonavir | Received within ≤5 days from diagnosis | 1,455 (18.9) | 6,239 (16.3) |
|  | Received >5 days from diagnosis | 25 (0.3) | 212 (0.6) |
|  | Not received | 6,214 (80.8) | 31,922 (83.2) |

<sup>1</sup>Counts and percentages are counted excluding missing values.

**Table S2:**
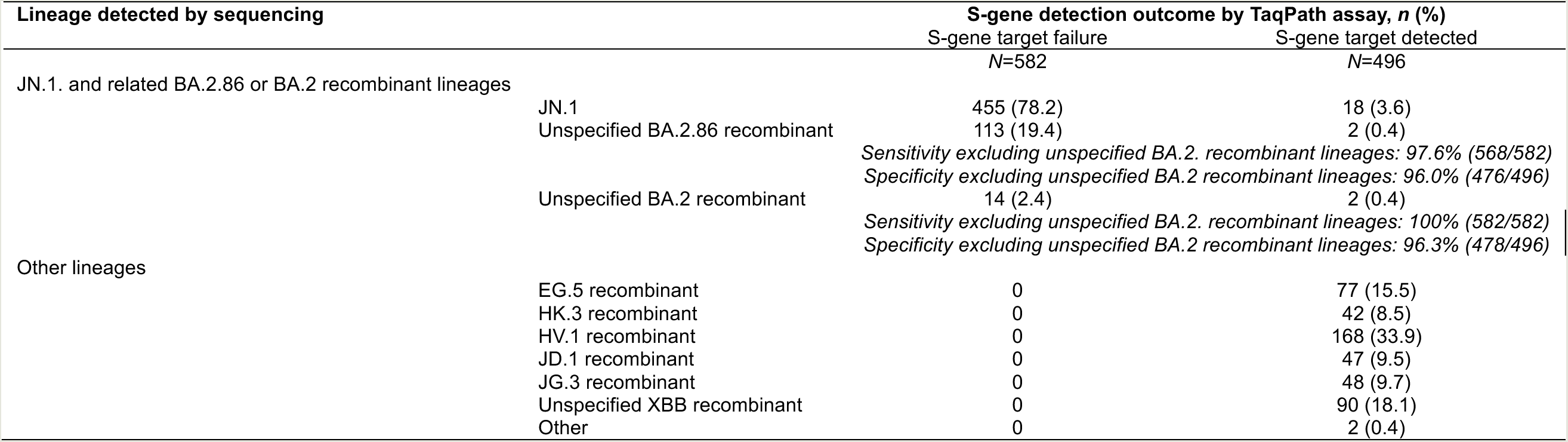
S-gene target detection and infecting lineage among outpatient-diagnosed cases with samples processed using the TaqPath COVID-19 Combo Kit assay.

**Table S3:**
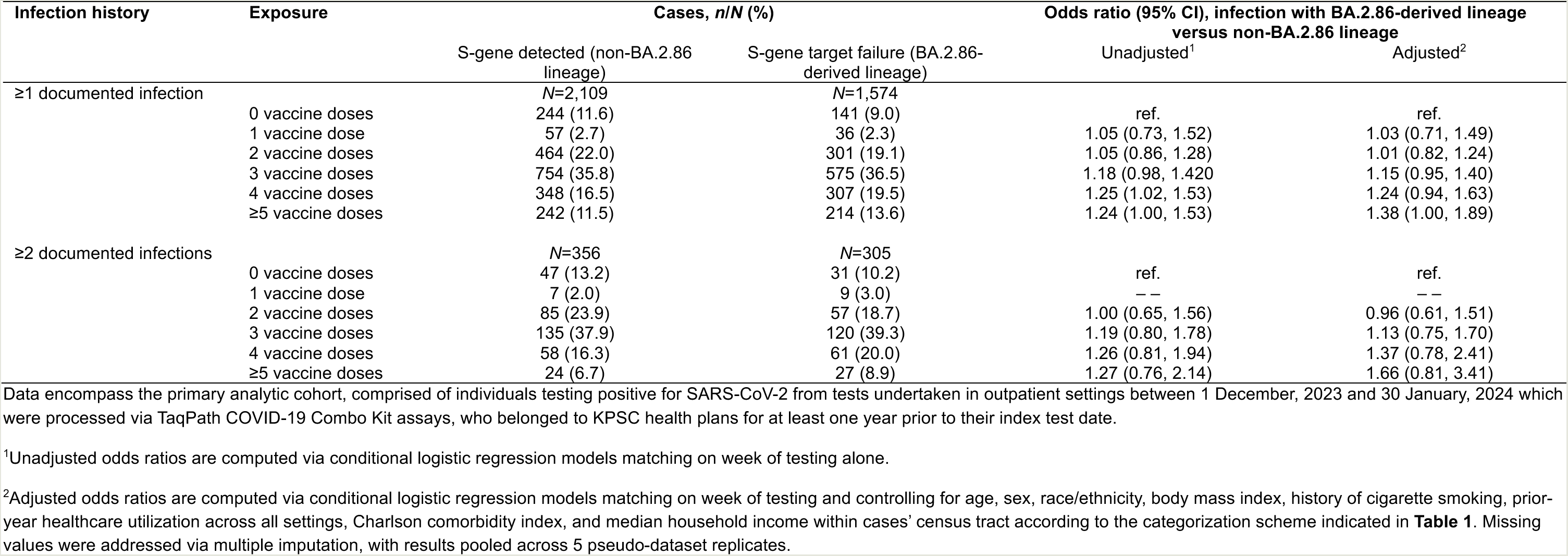
Prior vaccination and documented SARS-CoV-2 infection among cases infected with JN.1 and non-JN.1 lineages.

| Infection history | Exposure | Cases, *n/N* (%) |  | Odds ratio (95% CI), infection with BA.2.86-derived lineage versus non-BA.2.86 lineage |  |
| --- | --- | --- | --- | --- | --- |
|  |  | S-gene detected (non-BA.2.86 lineage) | S-gene target failure (BA.2.86-derived lineage) | Unadjusted<sup>1</sup> | Adjusted<sup>2</sup> |
| ≥1 documented infection |  | *N*=2,109 | *N*=1,574 |  |  |
|  | 0 vaccine doses | 244 (11.6) | 141 (9.0) | ref. | ref. |
|  | 1 vaccine dose | 57 (2.7) | 36 (2.3) | 1.05 (0.73, 1.52) | 1.03 (0.71, 1.49) |
|  | 2 vaccine doses | 464 (22.0) | 301 (19.1) | 1.05 (0.86, 1.28) | 1.01 (0.82, 1.24) |
|  | 3 vaccine doses | 754 (35.8) | 575 (36.5) | 1.18 (0.98, 1.420) | 1.15 (0.95, 1.40) |
|  | 4 vaccine doses | 348 (16.5) | 307 (19.5) | 1.25 (1.02, 1.53) | 1.24 (0.94, 1.63) |
|  | ≥5 vaccine doses | 242 (11.5) | 214 (13.6) | 1.24 (1.00, 1.53) | 1.38 (1.00, 1.89) |
| ≥2 documented infections |  | *N*=356 | *N*=305 |  |  |
|  | 0 vaccine doses | 47 (13.2) | 31 (10.2) | ref. | ref. |
|  | 1 vaccine dose | 7 (2.0) | 9 (3.0) | — | — |
|  | 2 vaccine doses | 85 (23.9) | 57 (18.7) | 1.00 (0.65, 1.56) | 0.96 (0.61, 1.51) |
|  | 3 vaccine doses | 135 (37.9) | 120 (39.3) | 1.19 (0.80, 1.78) | 1.13 (0.75, 1.70) |
|  | 4 vaccine doses | 58 (16.3) | 61 (20.0) | 1.26 (0.81, 1.94) | 1.37 (0.78, 2.41) |
|  | ≥5 vaccine doses | 24 (6.7) | 27 (8.9) | 1.27 (0.76, 2.14) | 1.66 (0.81, 3.41) |

Data encompass the primary analytic cohort, comprised of individuals testing positive for SARS-CoV-2 from tests undertaken in outpatient settings between 1 December, 2023 and 30 January, 2024 which were processed via TaqPath COVID-19 Combo Kit assays, who belonged to KPSC health plans for at least one year prior to their index test date.
<sup>1</sup>Unadjusted odds ratios are computed via conditional logistic regression models matching on week of testing alone.
<sup>2</sup>Adjusted odds ratios are computed via conditional logistic regression models matching on week of testing and controlling for age, sex, race/ethnicity, body mass index, history of cigarette smoking, prior-year healthcare utilization across all settings, Charlson comorbidity index, and median household income within cases' census tract according to the categorization scheme indicated in **Table 1**. Missing values were addressed via multiple imputation, with results pooled across 5 pseudo-dataset replicates.

**Table S4:** Putative prior infecting variants among individuals infected with SARS-CoV-2 according to lineage.

| Infection history | Cases, *n/N* (%) |  | Odds ratio (95% CI), infection with BA.2.86-derived lineage versus non-BA.2.86 lineage |  |
| --- | --- | --- | --- | --- |
|  | S-gene detected (non-BA.2.86 lineage)<br>*N*=4,614 | S-gene target failure (BA.2.86-derived lineage)<br>*N*=3,080 | Unadjusted<sup>1</sup> | Adjusted |
| 0 documented infections | 2,505 (54.3) | 1,506 (48.9) | ref. | ref. |
| Prior documented infection during pre-Delta period<sup>1</sup> | 572 (12.4) | 378 (12.3) | 1.00 (0.90, 1.12) | 1.00 (0.89, 1.12) |
| Prior documented infection during Delta period<sup>2</sup> | 155 (3.4) | 102 (3.3) | 0.97 (0.80, 1.19) | 0.95 (0.77, 1.16) |
| Prior documented infection during BA.1 period<sup>3</sup> | 621 (13.5) | 465 (15.1) | 1.08 (0.98, 1.20) | 1.08 (0.97, 1.19) |
| Prior documented infection during BA.2 period<sup>4</sup> | 303 (6.6) | 267 (8.7) | 1.17 (1.03, 1.33) | 1.16 (1.02, 1.32) |
| Prior documented infection during BA.4/BA.5 period<sup>5</sup> | 559 (12.1) | 414 (13.4) | 1.07 (0.96, 1.18) | 0.98 (0.87, 1.11) |
| Prior documented infection during XBB period<sup>6</sup> | 269 (5.8) | 264 (8.6) | 1.18 (1.04, 1.34) | 1.16 (1.02, 1.32) |

Data encompass the primary analytic cohort, comprised of individuals testing positive for SARS-CoV-2 from tests undertaken in outpatient settings between 1 December, 2023 and 30 January, 2024 which were processed via TaqPath COVID-19 Combo Kit assays, who belonged to KPSC health plans for at least one year prior to their index test date.
<sup>1</sup>Unadjusted odds ratios are computed via conditional logistic regression models matching on week of testing alone.
<sup>2</sup>Adjusted odds ratios are computed via conditional logistic regression models matching on week of testing and controlling for age, sex, race/ethnicity, body mass index, history of cigarette smoking, prior-year healthcare utilization across all settings, Charlson comorbidity index, and median household income within cases' census tract according to the categorization scheme indicated in **Table 1**. Missing values were addressed via multiple imputation, with results pooled across 5 pseudo-dataset replicates.
<sup>3</sup>Pre-Delta period: 1 January, 2020 to 19 June, 2021.
<sup>4</sup>Delta period: 20 June, 2021 to 19 December, 2021.
<sup>5</sup>BA.1 period: 20 December, 2021 to 2 February, 2022.
<sup>6</sup>BA.2 period: 3 February, 2022 to 24 June, 2022.
<sup>7</sup>BA.4/BA.5 period: 25 June, 2022 to 30 November, 2022.
<sup>8</sup>XBB period: 1 December, 2022 to 31 October, 2023.

**Table S5:**
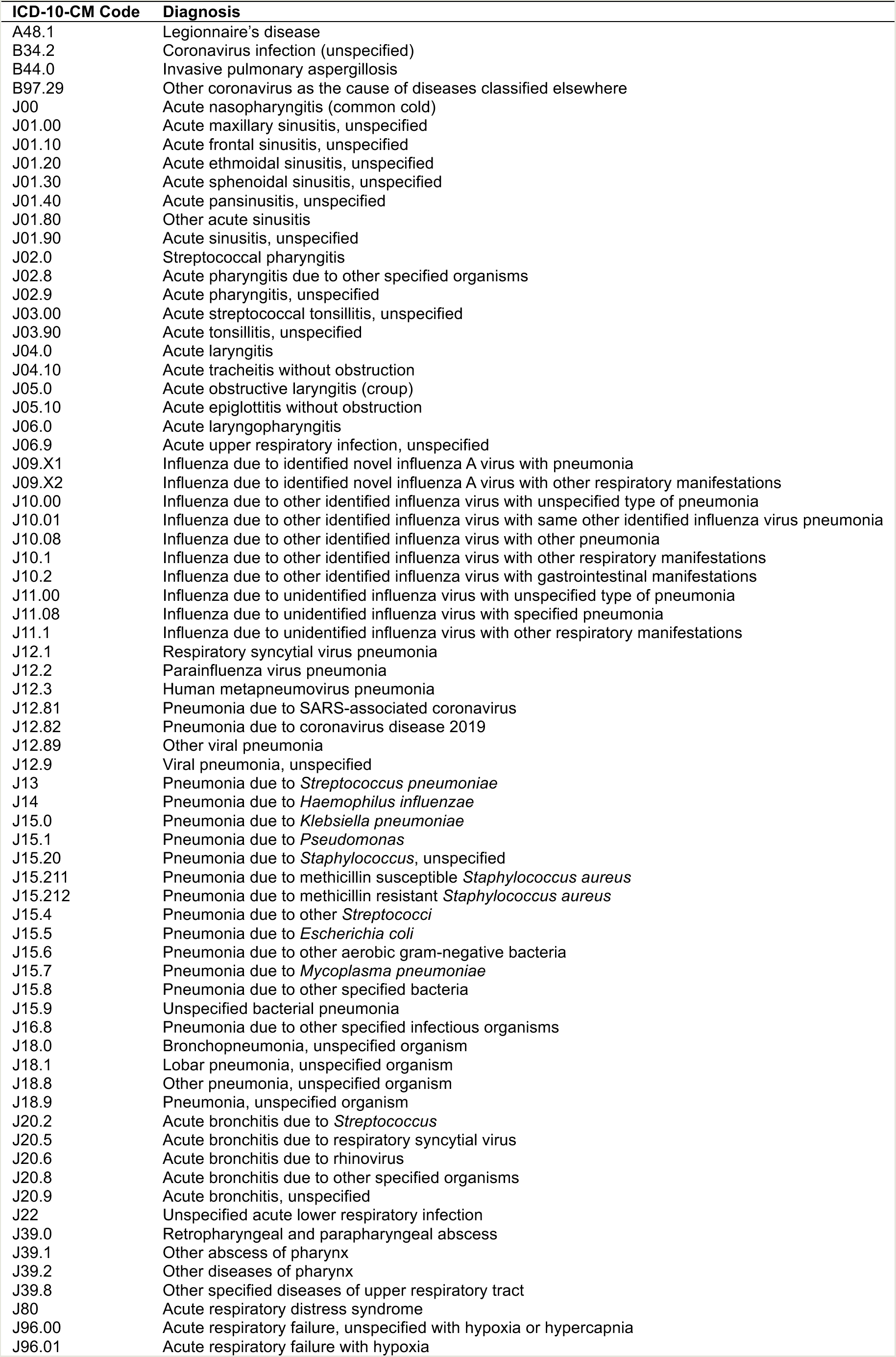
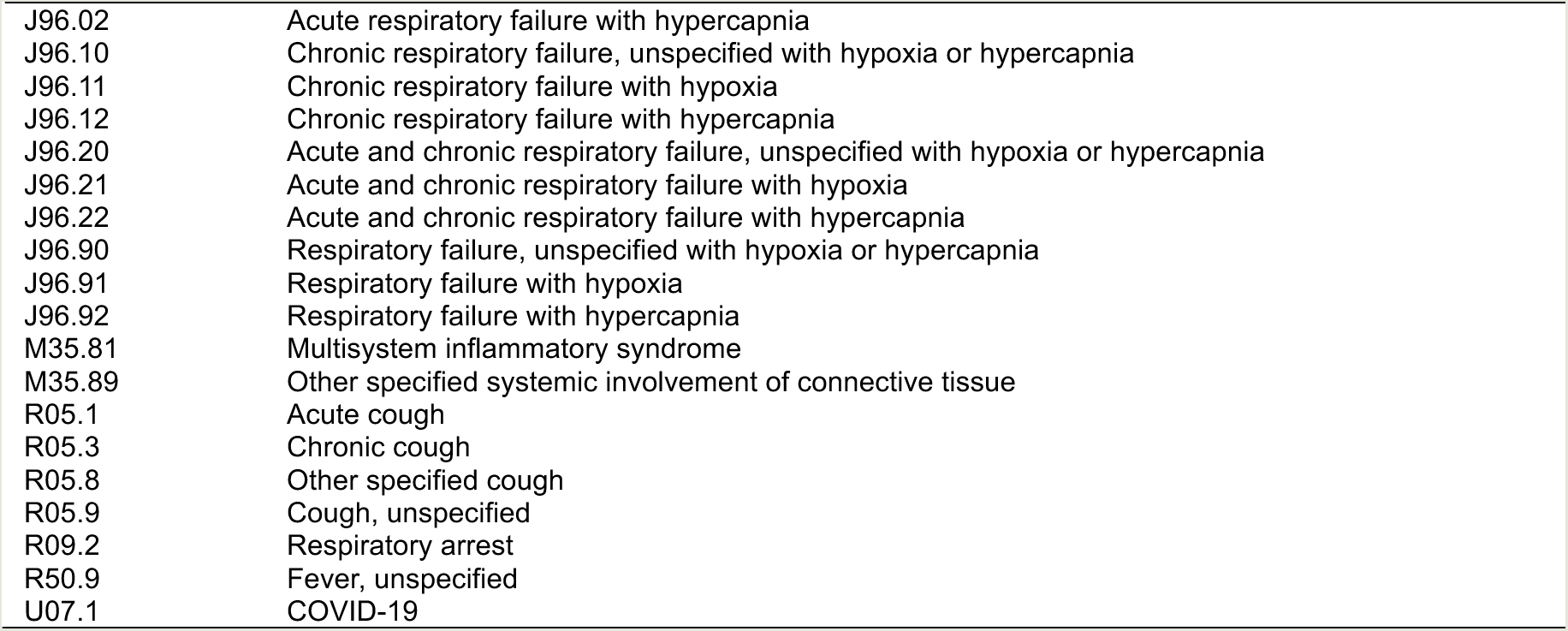
Diagnosis codes used to identify acute respiratory infection-associated emergency department presentations and hospital admissions.

| ICD-10-CM Code | Diagnosis |
| --- | --- |
| A48.1 | Legionnaire's disease |
| B34.2 | Coronavirus infection (unspecified) |
| B44.0 | Invasive pulmonary aspergillosis |
| B97.29 | Other coronavirus as the cause of diseases classified elsewhere |
| J00 | Acute nasopharyngitis (common cold) |
| J01.00 | Acute maxillary sinusitis, unspecified |
| J01.10 | Acute frontal sinusitis, unspecified |
| J01.20 | Acute ethmoidal sinusitis, unspecified |
| J01.30 | Acute sphenoidal sinusitis, unspecified |
| J01.40 | Acute pansinusitis, unspecified |
| J01.80 | Other acute sinusitis |
| J01.90 | Acute sinusitis, unspecified |
| J02.0 | Streptococcal pharyngitis |
| J02.8 | Acute pharyngitis due to other specified organisms |
| J02.9 | Acute pharyngitis, unspecified |
| J03.00 | Acute streptococcal tonsillitis, unspecified |
| J03.90 | Acute tonsillitis, unspecified |
| J04.0 | Acute laryngitis |
| J04.10 | Acute tracheitis without obstruction |
| J05.0 | Acute obstructive laryngitis (croup) |
| J05.10 | Acute epiglottitis without obstruction |
| J06.0 | Acute laryngopharyngitis |
| J06.9 | Acute upper respiratory infection, unspecified |
| J09.X1 | Influenza due to identified novel influenza A virus with pneumonia |
| J09.X2 | Influenza due to identified novel influenza A virus with other respiratory manifestations |
| J10.00 | Influenza due to other identified influenza virus with unspecified type of pneumonia |
| J10.01 | Influenza due to other identified influenza virus with same other identified influenza virus pneumonia |
| J10.08 | Influenza due to other identified influenza virus with other pneumonia |
| J10.1 | Influenza due to other identified influenza virus with other respiratory manifestations |
| J10.2 | Influenza due to other identified influenza virus with gastrointestinal manifestations |
| J11.00 | Influenza due to unidentified influenza virus with unspecified type of pneumonia |
| J11.08 | Influenza due to unidentified influenza virus with specified pneumonia |
| J11.1 | Influenza due to unidentified influenza virus with other respiratory manifestations |
| J12.1 | Respiratory syncytial virus pneumonia |
| J12.2 | Parainfluenza virus pneumonia |
| J12.3 | Human metapneumovirus pneumonia |
| J12.81 | Pneumonia due to SARS-associated coronavirus |
| J12.82 | Pneumonia due to coronavirus disease 2019 |
| J12.89 | Other viral pneumonia |
| J12.9 | Viral pneumonia, unspecified |
| J13 | Pneumonia due to *Streptococcus pneumoniae* |
| J14 | Pneumonia due to *Haemophilus influenzae* |
| J15.0 | Pneumonia due to *Klebsiella pneumoniae* |
| J15.1 | Pneumonia due to *Pseudomonas* |
| J15.20 | Pneumonia due to *Staphylococcus*, unspecified |
| J15.211 | Pneumonia due to methicillin susceptible *Staphylococcus aureus* |
| J15.212 | Pneumonia due to methicillin resistant *Staphylococcus aureus* |
| J15.4 | Pneumonia due to other *Streptococci* |
| J15.5 | Pneumonia due to *Escherichia coli* |
| J15.6 | Pneumonia due to other aerobic gram-negative bacteria |
| J15.7 | Pneumonia due to *Mycoplasma pneumoniae* |
| J15.8 | Pneumonia due to other specified bacteria |
| J15.9 | Unspecified bacterial pneumonia |
| J16.8 | Pneumonia due to other specified infectious organisms |
| J18.0 | Bronchopneumonia, unspecified organism |
| J18.1 | Lobar pneumonia, unspecified organism |
| J18.8 | Other pneumonia, unspecified organism |
| J18.9 | Pneumonia, unspecified organism |
| J20.2 | Acute bronchitis due to *Streptococcus* |
| J20.5 | Acute bronchitis due to respiratory syncytial virus |
| J20.6 | Acute bronchitis due to rhinovirus |
| J20.8 | Acute bronchitis due to other specified organisms |
| J20.9 | Acute bronchitis, unspecified |
| J22 | Unspecified acute lower respiratory infection |
| J39.0 | Retropharyngeal and parapharyngeal abscess |
| J39.1 | Other abscess of pharynx |
| J39.2 | Other diseases of pharynx |
| J39.8 | Other specified diseases of upper respiratory tract |
| J80 | Acute respiratory distress syndrome |
| J96.00 | Acute respiratory failure, unspecified with hypoxia or hypercapnia |
| J96.01 | Acute respiratory failure with hypoxia |
| J96.02 | Acute respiratory failure with hypercapnia |
| J96.10 | Chronic respiratory failure, unspecified with hypoxia or hypercapnia |
| J96.11 | Chronic respiratory failure with hypoxia |
| J96.12 | Chronic respiratory failure with hypercapnia |
| J96.20 | Acute and chronic respiratory failure, unspecified with hypoxia or hypercapnia |
| J96.21 | Acute and chronic respiratory failure with hypoxia |
| J96.22 | Acute and chronic respiratory failure with hypercapnia |
| J96.90 | Respiratory failure, unspecified with hypoxia or hypercapnia |
| J96.91 | Respiratory failure with hypoxia |
| J96.92 | Respiratory failure with hypercapnia |
| M35.81 | Multisystem inflammatory syndrome |
| M35.89 | Other specified systemic involvement of connective tissue |
| R05.1 | Acute cough |
| R05.3 | Chronic cough |
| R05.8 | Other specified cough |
| R05.9 | Cough, unspecified |
| R09.2 | Respiratory arrest |
| R50.9 | Fever, unspecified |
| U07.1 | COVID-19 |

**Table S6:**
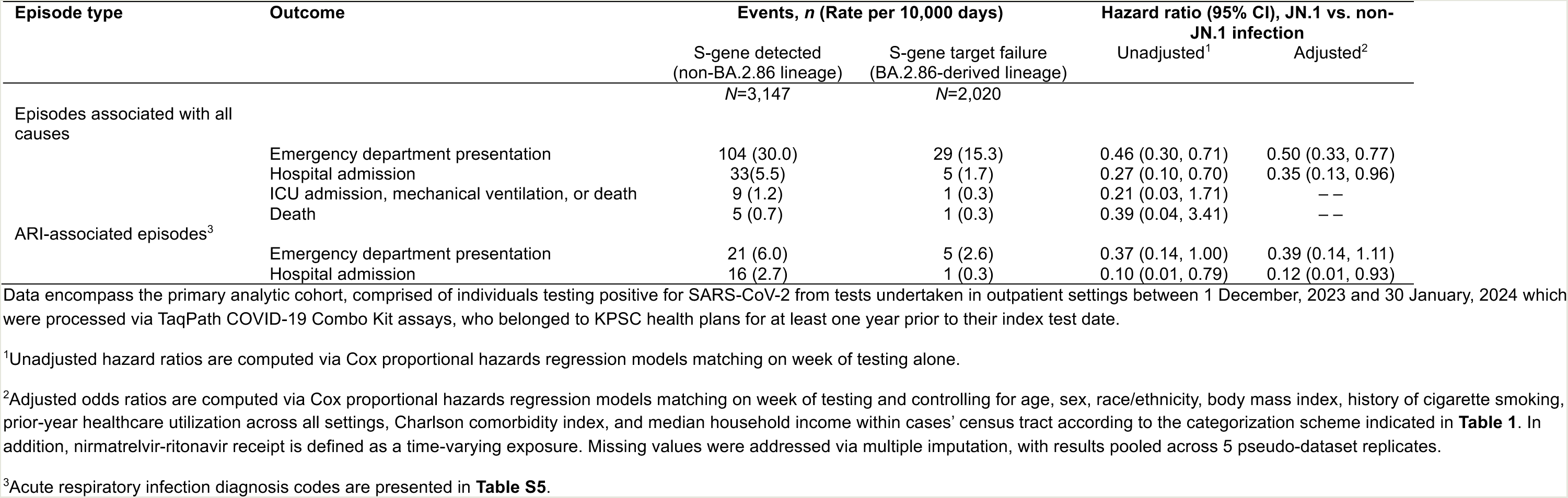
Clinical progression among cases according to infecting lineage, restricting the sample to cases with >5 healthcare interactions in the year preceding their index test.

| Episode type | Outcome | Events, *n* (Rate per 10,000 days) |  | Hazard ratio (95% CI), JN.1 vs. non-JN.1 infection |  |
| --- | --- | --- | --- | --- | --- |
|  |  | S-gene detected<br>(non-BA.2.86 lineage)<br>*N*=3,147 | S-gene target failure<br>(BA.2.86-derived lineage)<br>*N*=2,020 | Unadjusted<sup>1</sup> | Adjusted<sup>2</sup> |
| Episodes associated with all causes | Emergency department presentation | 104 (30.0) | 29 (15.3) | 0.46 (0.30, 0.71) | 0.50 (0.33, 0.77) |
|  | Hospital admission | 33(5.5) | 5 (1.7) | 0.27 (0.10, 0.70) | 0.35 (0.13, 0.96) |
|  | ICU admission, mechanical ventilation, or death | 9 (1.2) | 1 (0.3) | 0.21 (0.03, 1.71) | — |
|  | Death | 5 (0.7) | 1 (0.3) | 0.39 (0.04, 3.41) | — |
| ARI-associated episodes<sup>3</sup> | Emergency department presentation | 21 (6.0) | 5 (2.6) | 0.37 (0.14, 1.00) | 0.39 (0.14, 1.11) |
|  | Hospital admission | 16 (2.7) | 1 (0.3) | 0.10 (0.01, 0.79) | 0.12 (0.01, 0.93) |

Data encompass the primary analytic cohort, comprised of individuals testing positive for SARS-CoV-2 from tests undertaken in outpatient settings between 1 December, 2023 and 30 January, 2024 which were processed via TaqPath COVID-19 Combo Kit assays, who belonged to KPSC health plans for at least one year prior to their index test date.
<sup>1</sup>Unadjusted hazard ratios are computed via Cox proportional hazards regression models matching on week of testing alone.
<sup>2</sup>Adjusted odds ratios are computed via Cox proportional hazards regression models matching on week of testing and controlling for age, sex, race/ethnicity, body mass index, history of cigarette smoking, prior-year healthcare utilization across all settings, Charlson comorbidity index, and median household income within cases' census tract according to the categorization scheme indicated in **Table 1**. In addition, nirmatrelvir-ritonavir receipt is defined as a time-varying exposure. Missing values were addressed via multiple imputation, with results pooled across 5 pseudo-dataset replicates.
<sup>3</sup>Acute respiratory infection diagnosis codes are presented in **Table S5**.

**Figure S1:**
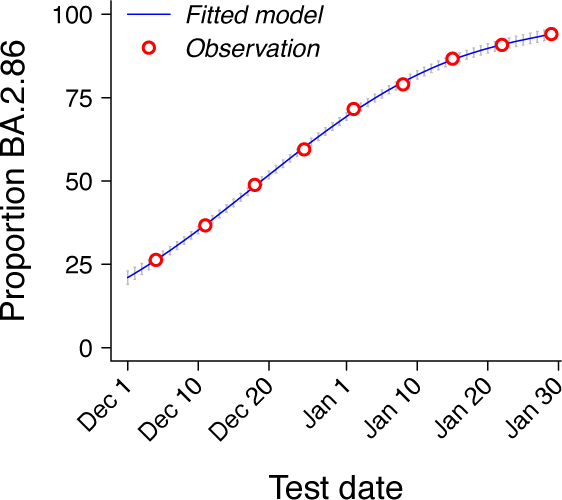
Observed and fitted proportions of cases infected with BA.2.86 lineages. We illustrate weekly proportions of cases found to be infected with BA.2.86 lineages (points, plotted at weekly mid-points), based on sequencing of a random selection of cases across all test settings, against model fitted with a 5^th^-degree polynomial function. The blue center line corresponds to median estimates, while grey vertical bars delineate 95% confidence intervals in day-specific predictions.

